# Urban environment and socio-economic inequalities in childhood excess weight: a cross-sectional study in Geneva, Switzerland

**DOI:** 10.64898/2026.05.26.26354079

**Authors:** Viviane Richard, David De Ridder, Harris Héritier, Elsa Lorthe, Roxane Dumont, Nicolas Bovio, Mayssam Nehme, Rémy P. Barbe, Klara M. Posfay-Barbe, Thomas W. McDade, Nicolas Vuilleumier, Idris Guessous, Silvia Stringhini, SEROCoV-KIDS study group

## Abstract

**Background:** Childhood overweight and obesity represent major public health challenges, shaped by socio-economic and environmental factors. This study investigates the mediating and moderating role of urban environmental exposures in socio-economic disparities in childhood excess weight.

**Methods:** Data was drawn from a population-based sample of children (2-9 years) and adolescents (10-17 years) living in Geneva, Switzerland. Parents reported household financial situation and children’s height and weight, from which excess weight (i.e. overweight or obesity) was derived. Residential exposures to air pollution (PM2.5, NO_2_), noise (daytime, nighttime), and neighborhood greenness (green areas, canopy coverage) were estimated based on geocoded residential addresses. The association between household financial situation and excess weight was evaluated, as well as the mediating and moderating roles of urban environmental exposures.

**Results:** The analysis included 1’006 children and 1’154 adolescents. Among children, an average-to-poor household financial situation was associated with higher odds of excess weight in children (adjusted odds ratio [aOR]: 1.79, 95% confidence interval [CI]: 1.13; 2.84). Higher noise exposure was associated with excess weight (daytime: aOR: 1.40, 95% CI: 1.10; 1.77, nighttime: aOR: 1.37, 95% CI: 1.08; 1.74), while the association with PM2.5 appeared stronger among socio-economically disadvantaged children, though the interaction did not reach statistical significance (financial situation × PM2.5 interaction: aOR: 1.59, 95% CI: 0.98; 2.59). No significant associations were observed among adolescents.

**Conclusion:** These findings highlight the joint influence of social and environmental inequalities on childhood excess weight and stress the need to address these interconnected determinants to design equitable, targeted public health interventions.

## 1. INTRODUCTION

Childhood overweight and obesity affect one in five children and adolescents worldwide [1], representing major public health concerns as they constitute risk factors for cardiometabolic conditions, including dyslipidemia, type 2 diabetes and hypertension [2]. In high-income countries, excess weight is more prevalent among children from lower socio-economic backgrounds [3], [4]. Urban environmental conditions, such as elevated air and noise pollution and limited access to green spaces, are associated with excess weight and may contribute to these socio-economic inequalities [5].

Exposure to outdoor air pollution has been linked to childhood overweight, potentially through chronic inflammation, oxidative stress, and reduced opportunities for physical activity [6], [7]. Evidence regarding noise is less consistent, yet transportation-related noise might also contribute to excess weight via increased stress and sleep disturbances [8], [9]. Conversely, green space and vegetation exposure has been associated with reduced weight [10], [11]. These benefits might arise from increased opportunities for physical activity and social cohesion but also from restorative effects and decreased stress [10]. Children are considered particularly vulnerable to such environmental influences due to ongoing physiological development, a higher air intake relative to body weight, and fewer coping strategies or autonomy to deal with disturbances than adults [5], [12].

Environmental effects on health outcomes, including weight outcomes, tend to be socio-economically patterned [13], [14]. Two non-exclusive mechanisms have been proposed: differential exposure and differential susceptibility [13], [14]. The *differential exposure* hypothesis posits that disadvantaged individuals are more likely to live in health-adverse environmental conditions, such as high air and noise pollution, and poorer access to quality green spaces, which are thus thought to *mediate* socio-economic inequalities in health [13]. Despite broad recognition of this pathway, few studies have quantified mediation for weight outcomes. A cross-country European study showed an indirect negative effect of household income on child body mass index (BMI) through neighboring green spaces that promoted engagement in physical activity [15]. On the contrary, limited access to a private garden or green spaces did not mediate the relationship between unfavorable socio-economic circumstances and overweight in United Kingdom children [16]. To the best of our knowledge, no study has quantified the mediating role of air and noise pollution in socio-economic inequalities in overweight.

The *differential susceptibility* hypothesis instead proposes that disadvantaged individuals experience more severe health impacts of equivalent exposures due to greater underlying vulnerability, implying a moderating effect [13]. Supporting this, a large longitudinal Spanish study found stronger associations between air pollution and overweight among children in more deprived areas [17]. Evidence regarding noise pollution and overweight is limited, though increased susceptibility to noise exposure in lower-income children has been reported for other outcomes, such as behavioral problems [18]. Conversely, green space access may confer greater protection against excess weight among disadvantaged children [16], [19], [20], although findings are mixed [21]. Overall, the complex pathways by which urban environmental exposures contribute to socio-economic disparities in children’s weight remain underexplored, especially for noise.

The current study was conducted in the canton of Geneva, a densely populated region of 246 km^2^ (2’134 inhabitants per km^2^) with a mix of urban and peri-urban environments [22]. Although Geneva ranks highly on the European Healthy Urban Design Index [23], average air pollution levels exceed World Health Organization guidelines in urban areas (WHO) [24], 37% of the cantonal population suffers from traffic-related noise disturbance [25], and socio-economic inequalities persist in the distribution of quality green spaces [26].

In this setting, the study investigates the mediating and moderating role of urban environmental exposures, including air and noise pollution and greenness, in socio-economic disparities in childhood excess weight.

## 2. METHODS

### 2.1. Study design

Data were drawn from the baseline assessment of the SEROCoV-KIDS prospective cohort study and a seroprevalence study with similar recruitment and data collection methods [27], [28]. Children were eligible to participate if aged between 6 months and 17 years and residing in the canton of Geneva, Switzerland. Participants were either randomly selected from official state registries (or had a sibling selected) or belonged to a household participating in one of the previous population-based SARS-CoV-2 seroprevalence studies conducted by our team. All referent adults, as well as adolescents aged 14 years or older provided written consent to participate; children gave oral assent. The Geneva Cantonal Commission for Research Ethics approved the seroprevalence study (ID: 2020–00881), the SEROCoV-KIDS study (ID: 2021-01973), and the CIFAR study for additional analysis of environmental exposure on existing data (ID: 2024-02567).

At enrollment, one referent parent per household completed a household socio-demographic questionnaire and a health questionnaire for each participating child. Recruitment and data collection took place between December 2021 and June 2022.

### 2.2. Study population

The analysis included children aged 2 years and older at baseline (n=2’188). Participants whose reported place of residence was outside the canton of Geneva were excluded (n=24). Four children with a sex reported as “other” were additionally excluded because the small number in this category precluded model estimation. The final analytical sample comprised a total of 2160 individuals, divided into 1’006 children aged 2-9 years and 1’154 adolescents aged 10-17 years.

### 2.3. Conceptual model

Variable choice and model specifications were guided by hypothesized relationships between study variables, summarized in a directed acyclic graph (DAG, Figure 1). The first research question (Q1) addressed the association between socio-economic conditions and excess weight. Although we acknowledge the key role of diet, physical activity and sleep in excess weight, they were not included in the model. Indeed, as potential mediators [29], including them would attenuate the total effect of socio-economic conditions, which was the estimate of interest for Q1. In a second step, the mediating (Q2a & Q2b) and moderating (Q3) role of environmental exposures in this association was assessed. As sleep duration and physical activity are hypothesized to mediate the environmental exposure - excess weight relationship [8]–[10], their contribution was explored when a significant total effect of environmental exposure on excess weight was observed. Diet was not examined as a mediator of environmental effects because of lower theoretical relevance and because detailed dietary data were not collected.

**Figure 1.**
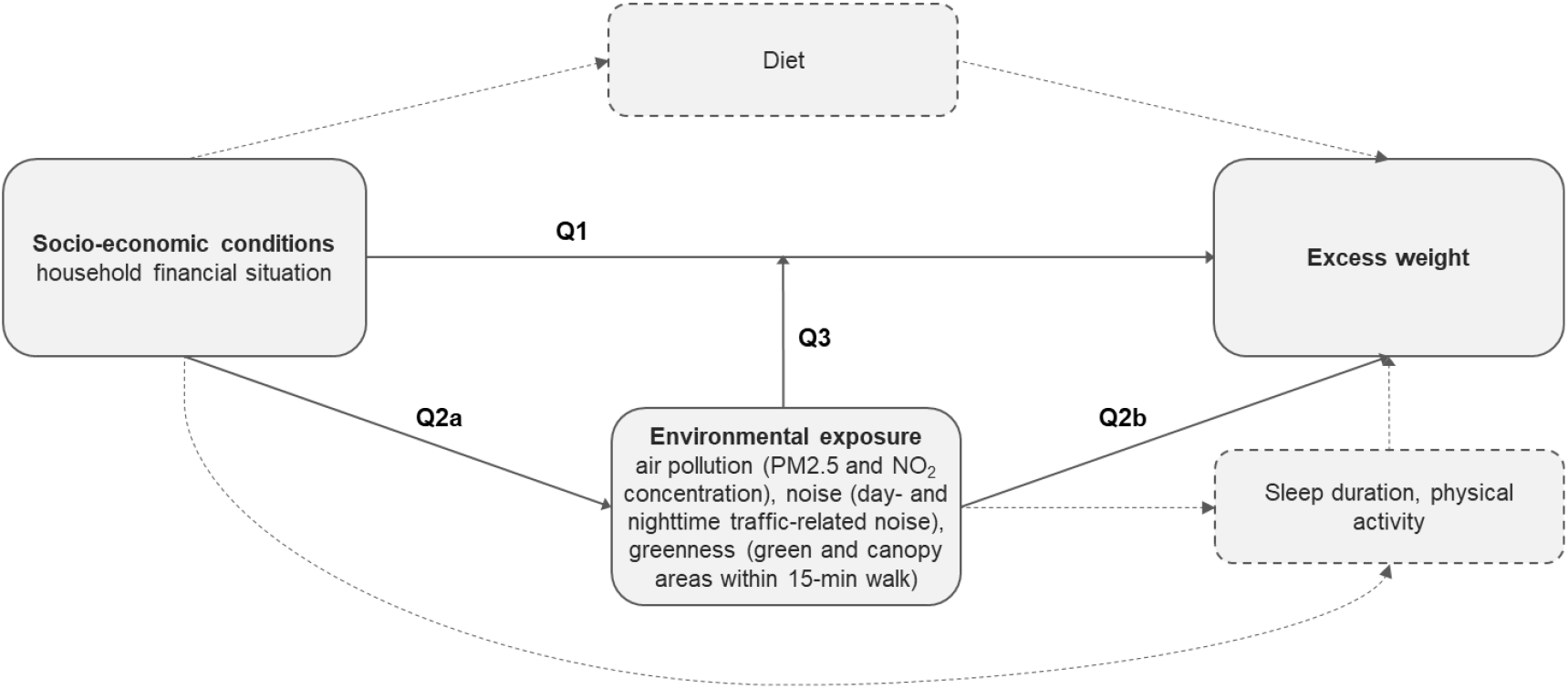
Directed acyclic graph (DAG) of the hypothesized relationships between study variables and corresponding research questions (Q). Solid arrows represent the main associations examined in the present study; dashed arrows indicate relevant pathways that are not the primary focus of the analysis. Age, sex, and parents’ birth country are covariates of all associations but are omitted for readability. Socio-economic conditions, environmental exposures, and parents’ birth country are measured at the household level.

### 2.4. Measures

#### 2.4.1. Health outcome

Parents were asked to measure their children’s height and weight specifically for this study, following detailed instructions. Age- and sex-specific z-scores were derived from these measures. Z-scores exceeding the 2024 United States Center for Disease and Control and Prevention (CDC) plausibility thresholds were considered implausible and set as missing (n=17) : weight <−5 or >8 standard deviations [SD], height <−5 or >4 SDs) or BMI <−4 or >8 SDs [30]. Excess weight, considered as a binary variable, was defined as BMI z-scores corresponding to overweight or obesity according to the WHO thresholds, i.e. above 2 SDs for children aged below 5 years and above 1 SD for older children and adolescents [31].

#### 2.4.2. Socio-economic exposure

Household financial situation was assessed by a single survey item. Responses were categorized as *good* if the referent parent reported having enough resources to live comfortably and save money or if they could meet their need and face small unexpected expenses, and *average to poor* if small unexpected expenses could cause financial difficulties or if they had to rely on regular external financial support. Participants who stated that they did not wish to provide this information were coded as missing.

#### 2.4.3. Urban environment exposures

Outdoor air pollution exposure was characterized using two pollutants primarily emitted by motorized traffic: particulate matter of less than 2.5 microns (PM2.5) and nitrogen dioxide (NO_2_). Raster data in a 100-meter and 20-meter grid, respectively, were obtained from the Swiss Federal Spatial Data Infrastructure [32]. For each pollutant, the average concentration of 2019-2023 values (µg/m^3^) within a 20-meter buffer around participants’ residences was calculated. Multi-year averages were used to characterize long-term residential exposure, assuming relative spatial stability in pollution patterns

Transport-related noise exposure data for road, rail and aircraft were obtained from the Swiss Federal Spatial Data Infrastructure [32]. Road and rail data were from 2021; aircraft data were from 2025. Equivalent continuous sound pressure levels (dB [A]) were provided for each transportation mode and combined to compute total transport-related noise exposure during daytime (6a.m. - 10p.m.) and nighttime (10p.m. - 6a.m.) with a 10×10-meter resolution. Day- and nighttime noise levels were analyzed separately to capture the potentially greater health impact of nighttime noise [33]. Total transport-related noise exposure was assigned to each participant based on residential location.

Greenness exposure was assessed by two complementary indicators: total green area (reflecting opportunities for active use) and tree canopy coverage (reflecting passive exposure) surrounding participants’ residential address, respectively. Green areas were derived from 2024 land cover data and included public and private forests, lawns, fields, wetlands, and vineyards [32]. Canopy coverage was derived from a 2023 cantonal model [34]. Both indicators were calculated within a 15-minute walking isochrone from each participant’s residence, based on the street network estimated using the R osrm package [35].

#### 2.4.4. Health behaviors

Sleep duration and physical activity (from 3 years old) were reported in hours per day for weekdays and weekends. For both behaviors, the average daily time was calculated as: (week time × 5 + weekend time × 2) / 7 and used as a continuous variable.

#### 2.4.5. Covariates

Children’s sex and age at inclusion, as well as parents’ birth country (*At least one born in Switzerland* vs *Both born abroad*) were collected in socio-demographic questionnaires.

### 2.5. Statistical analysis

The association between the household financial situation and excess weight (Q1, Figure 1) was estimated with age-, sex-, and parents’ birth country-adjusted generalised estimating equations (GEE) to account for the household clustering of data.

The differential exposure hypothesis was examined with two sets of GEE models separately estimating the associations between the household financial situation and each standardized urban environmental exposure, adjusted for age, sex, and parents’ birth country (Q2a, Figure 1), and the associations between each standardized urban environmental exposure and excess weight (Q2b, Figure 1). In the second set, minimally adjusted models controlled for age, sex, parents’ birth country and financial situation, while fully adjusted models additionally accounted for environmental exposures from other sources (e.g. nighttime noise - excess weight association adjusted for age, sex, parents’ birth country, financial situation, PM2.5, NO_2_, and green and canopy areas). When both Q2a and Q2b were statistically significant at a 5% level for a given environmental exposure, a formal counterfactual mediation analysis was conducted, adjusting for age, sex, parents’ birth country and environmental exposures from other sources.

The differential susceptibility hypothesis (Q3, Figure 1) was tested by separately adding an interaction term between the household financial situation and each urban environmental exposure to the fully adjusted models assessing the association between environmental exposures and excess weight. Models with excess weight as the outcome (Q1, Q2b & Q3, Figure 1) were stratified by age group (children: 2-9 years; adolescents: 10-17 years) to assess age-related patterns.

In exploratory analysis, when a given environmental exposure was significantly associated with excess weight at a 5% level (Q2b), the mediating role of sleep duration (for noise pollution) and physical activity (for air pollution and greenness) was assessed. Analyses followed a regression-based approach and were adjusted for age, sex, parents’ birth country, household financial situation, and environmental exposures from other sources. Analysis was based on complete case.

## 3. RESULTS

A total of 2’160 participants (1’006 children and 1’154 adolescents) were included with a mean age of 9.9 years (SD: 4.2), 49.4% being female (Table 1). Overall, 158 children (16.4%) and 189 adolescents (16.4%) had excess weight. Mean PM2.5 and NO_2_ concentration were 9.5 and 16.9 µg/m^3^, respectively, while day- and nighttime noise averaged 48.5 and 38.2 dB [A], respectively. Within a 15-minute walking distance from participants’ residences, the mean canopy area was 0.4 km^2^ and the mean green space area was 0.6 km^2^. Air pollution, noise and canopy areas were positively correlated with one another and negatively correlated with green areas (P-value < 0.001, Supplementary figure 1). Overall, 167 participants (7.7%) had at least one missing value on main study variables, primarily due to non-disclosure of household financial situation and implausible anthropometric values (Table 1). Those were excluded from corresponding analyses.

**Table 1.** Participants’ demographic, socio-economic and environmental by weight status.

|  | Children |  |  |  | Adolescents |  |  |  |
| --- | --- | --- | --- | --- | --- | --- | --- | --- |
|  | All<br>(n=1'006)<sup>a</sup> | Normal<br>weight<br>(n=829) | Excess<br>weight<br>(n=158) | p-value | All<br>(n=1154)<sup>a</sup> | Normal<br>weight<br>(n=964) | Excess<br>weight<br>(n=189) | p-value |
| **Age (years)** | 6.10 (2.24) | 5.95 (2.26) | 7.05 (1.82) | <0.001 | 13.12 (2.26) | 13.25 (2.28) | 12.43 (2.05) | <0.001 |
| **Sex** |  |  |  | 0.569 |  |  |  | 0.251 |
| Male | 526 (52.3) | 435 (52.5) | 79 (50.0) |  | 567 (49.1) | 466 (48.3) | 100 (52.9) |  |
| Female | 480 (47.7) | 394 (47.5) | 79 (50.0) |  | 587 (50.9) | 498 (51.7) | 89 (47.1) |  |
| **Parents' birth country** |  |  |  | 0.063 |  |  |  | 0.040 |
| At least one born in Switzerland | 582 (57.9) | 491 (59.2) | 81 (51.3) |  | 718 (62.2) | 612 (63.5) | 105 (55.6) |  |
| Both born abroad | 424 (42.1) | 338 (40.8) | 77 (48.7) |  | 436 (37.8) | 352 (36.5) | 84 (44.4) |  |
| **Household financial situation** |  |  |  | 0.007 |  |  |  | 0.202 |
| Good | 777 (82.9) | 661 (84.7) | 108 (75.5) |  | 851 (79.5) | 718 (80.1) | 132 (75.9) |  |
| Average to poor | 160 (17.1) | 119 (15.3) | 35 (24.5) |  | 220 (20.5) | 178 (19.9) | 42 (24.1) |  |
| Missing<sup>b</sup> | 69 | 49 | 15 |  | 83 | 68 | 15 |  |
| **PM2.5 concentration (µg/m<sup>3</sup>)** | 9.47 (0.59) | 9.46 (0.59) | 9.56 (0.58) | 0.084 | 9.45 (0.59) | 9.44 (0.58) | 9.52 (0.61) | 0.075 |
| **NO<sub>2</sub> concentration (µg/m<sup>3</sup>)** | 17.09 (3.09) | 17.02 (3.09) | 17.54 (3.13) | 0.099 | 16.79 (3.04) | 16.71 (3.02) | 17.24 (3.12) | 0.025 |
| **Daytime noise (dB[A])** | 48.50 (7.95) | 48.18 (7.94) | 50.24 (7.62) | 0.004 | 48.51 (8.07) | 48.28 (7.97) | 49.69 (8.49) | 0.012 |
| **Nighttime noise (dB[A])** | 38.24 (7.29) | 38.00 (7.29) | 39.60 (7.06) | 0.006 | 38.21 (7.35) | 38.02 (7.26) | 39.19 (7.77) | 0.024 |
| **Canopy areas within 15-min walk (km<sup>2</sup>)** | 0.38 (0.14) | 0.37 (0.14) | 0.39 (0.14) | 0.117 | 0.37 (0.13) | 0.37 (0.13) | 0.38 (0.13) | 0.466 |
| **Green areas within 15-min walk (km<sup>2</sup>)** | 0.54 (0.35) | 0.55 (0.36) | 0.50 (0.31) | 0.294 | 0.57 (0.34) | 0.58 (0.34) | 0.53 (0.32) | 0.069 |
| **Physical activity (hours/day)<sup>c</sup>** | 1.55 (0.88) | 1.56 (0.89) | 1.40 (0.80) | 0.029 | 1.24 (0.79) | 1.24 (0.79) | 1.20 (0.77) | 0.352 |
| **Sleep duration (hours/day)<sup>d</sup>** | 10.28 (1.15) | 10.35 (1.16) | 9.87 (0.93) | <0.001 | 8.97 (0.85) | 8.97 (0.85) | 8.93 (0.82) | 0.585 |

Results are numbers (%) and mean (standard deviation), p-values are from Chi<sup>2</sup> and Wilcoxon tests for categorical and continuous variables, respectively.
<sup>a</sup> 20 participants with missing excess weight, of which 17 because of implausible values <sup>b</sup> Missing for 152 participants, of which 147 did not wish to answer <sup>c</sup> Missing for 81 participants, including 78 children aged <3 years for whom the information was not collected <sup>d</sup> missing for 3 participants

Children from households with an average to poor financial situation had 1.79 times higher odds of having excess weight compared to their more privileged peers (adjusted odds ratio [aOR]: 1.79, 95% confidence interval [CI]: 1.13; 2.84), while there was no association in adolescents (aOR: 1.24, 95% CI: 0.82; 1.88, Supplementary table 1).

Looking at the differential exposure hypothesis, children and adolescents from disadvantaged households were more exposed to air pollution (standardized [std] β = 0.28, 95% CI: 0.13; 0.43 for PM2.5; std β = 0.25, 95% CI: 0.09; 0.40 for NO_2_) than their more advantaged peers (Supplementary table 1). They also had access to fewer green areas within a 15-minute walk (std β = −0.19, 95% CI: −0.33; −0.05), whereas no difference was observed in noise pollution and canopy coverage (p-value > 0.1).

Greater exposure to day- and nighttime noise was associated with a higher likelihood of excess weight among children, in both minimally and fully adjusted models (aOR: 1.40, 95% CI: 1.10; 1.77 for daytime; aOR: 1.37, 95% CI: 1.08; 1.74 for nighttime in fully adjusted models). Air pollution and greenness variables were not significantly related to excess weight (Figure 2).

**Figure 2.**
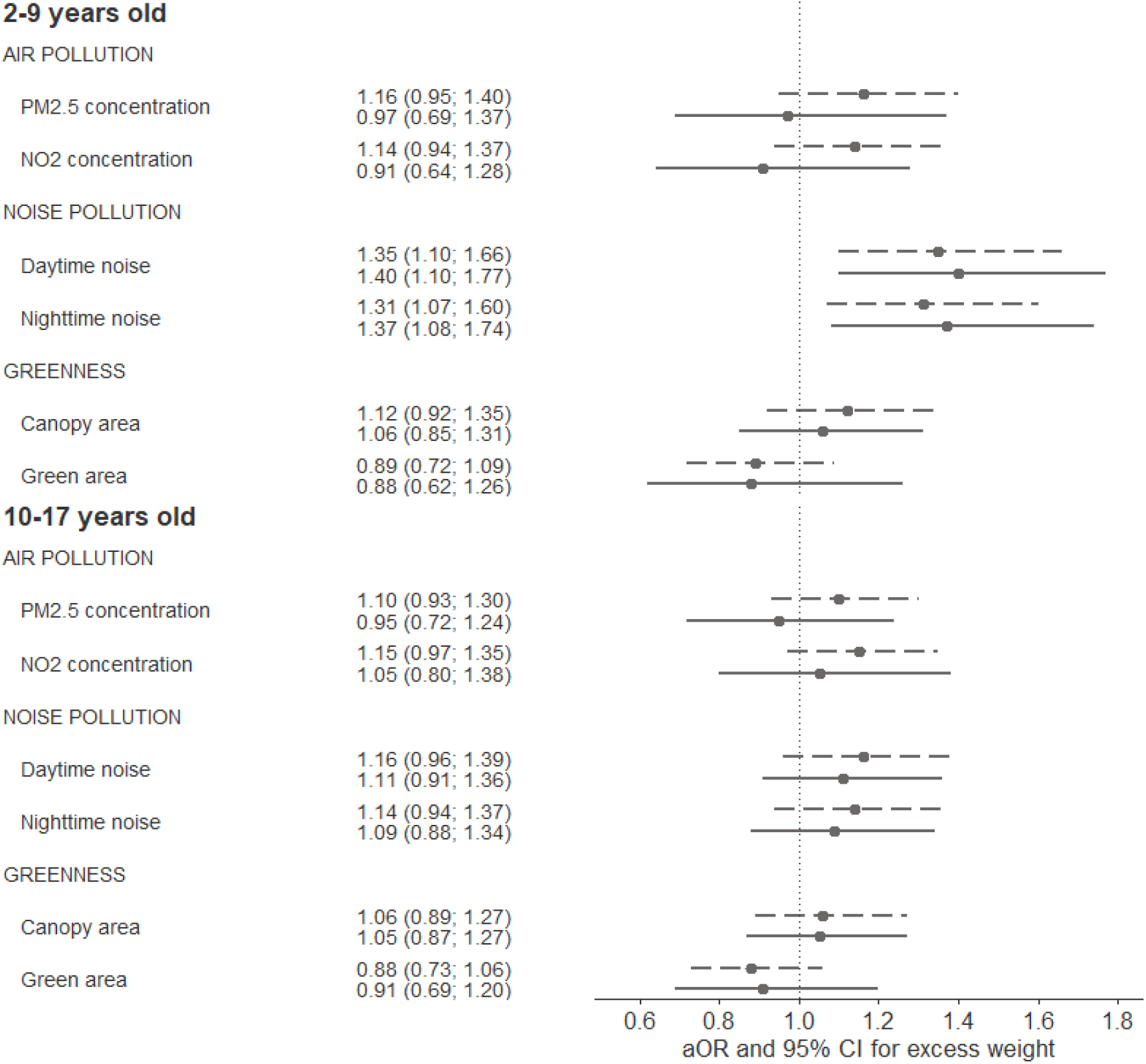
Associations between urban environmental exposures and excess weight in children (2-9 years old, n=923) and adolescents (10-17 years old, n=1’070) from generalized estimating equations. Results are adjusted odds ratios (aOR) and 95% confidence intervals (CI). Dashed lines correspond to the minimal models adjusted for age, sex, parents’ birth country and household financial situation; solid lines correspond to the full models additionally adjusted for types of environmental exposures not included as exposure (e.g. the association between canopy area and excess weight is adjusted for air and noise pollution measures). Urban environmental exposures are standardized; the corresponding coefficients express the odds of excess weight for a change of one standard deviation in the environmental exposure.

Although the household financial situation was associated with some urban environmental exposures (air pollution, green areas), it was not related to those associated with excess weight (noise pollution). Therefore, formal mediation analyses were not supported. Exploratory analyses examining sleep duration as a potential mediator of the association between noise pollution and excess weight showed no significant indirect effect (p-value > 0.1; Supplementary Table 2).

When examining the differential susceptibility hypothesis, the association of PM2.5 with excess weight appeared stronger, with a borderline significance level, in children with an average to poor household financial situation compared with their more privileged counterparts (aOR: 1.59, 95% CI: 0.98; 2.59, Figure 3). The interaction terms for other urban environmental exposures or among adolescents were non-significant and mostly centered around one (Supplementary table 3).

**Figure 3.**
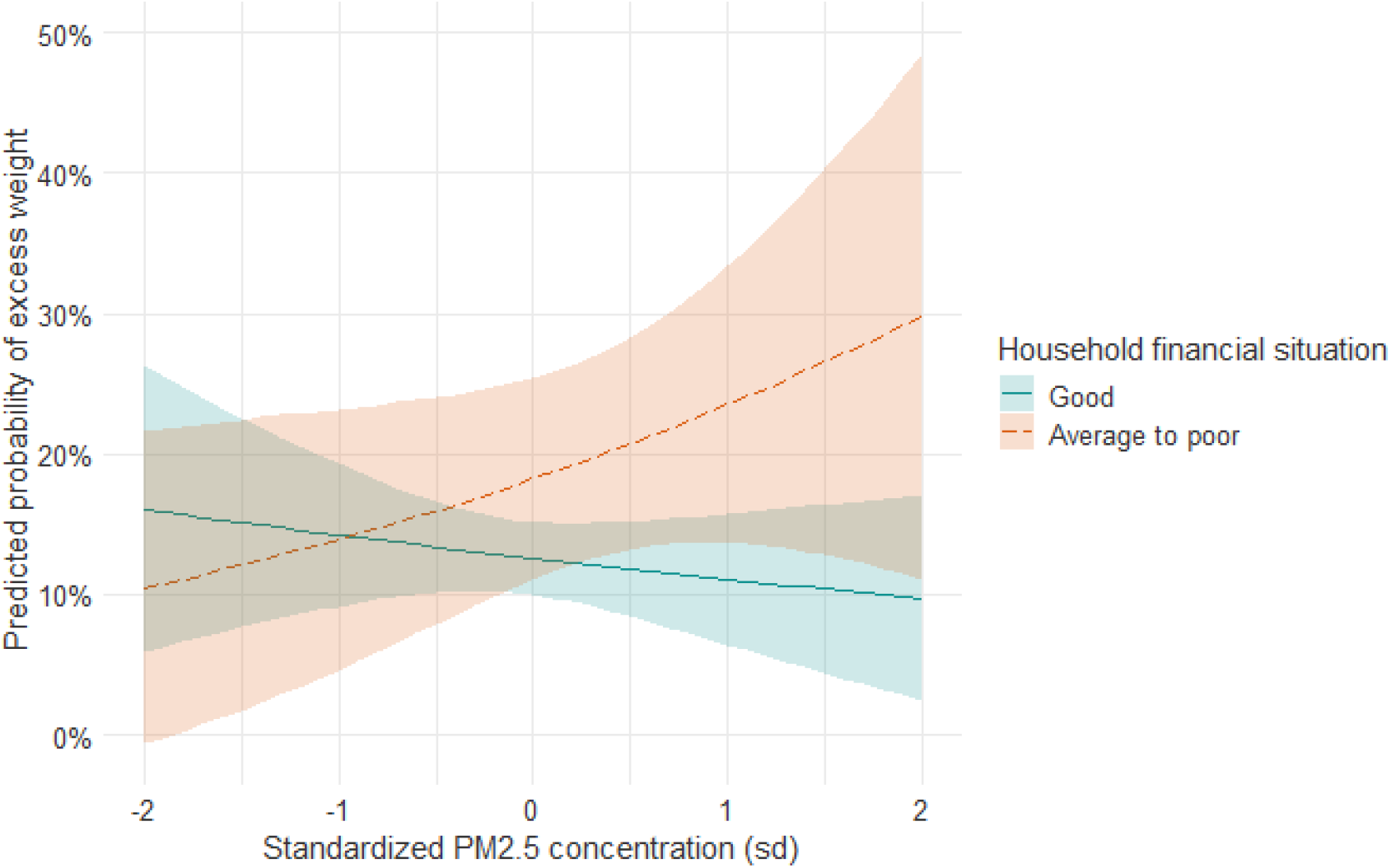
Predicted probability of excess weight by air pollution level in standard deviation (sd) and household financial situation in 2-9 years old children (n=923). Estimations from marginal means after generalized estimating equations adjusted for age, sex, parents’ birth country, noise pollution and greenness. P-value = 0.061.

## 4. DISCUSSION

In this study, an average to poor household financial situation was associated to both elevated likelihood of excess weight in children and to greater exposure to urban environmental stressors, such as air pollution and limited green areas. Noise pollution was associated with excess weight, while socio-economic disadvantage seemed to exacerbate the effect of air pollution.

Consistent with evidence from high-income countries, children from disadvantaged households were more likely to have excess weight [3], [4]. However, this pattern was not observed in adolescents, which contributes to the literature showing mixed evidence regarding the socio-economic gradient in overweight across age groups [3], [4]. Our findings may reflect an equalization mechanism in adolescence, whereby the influence of family decreases relative to peers [36]. This also aligns with previous results from the SEROCoV-KIDS cohort, which showed that socio-economic differences in health behaviors, such as screen time, physical activity, and sleep, decreased in adolescents [37].

As widely reported [13], socio-economic inequalities in the distribution of urban environmental exposures were also observed, with disadvantaged children being more likely to live in neighborhoods with higher air pollution and lower availability of green areas. These patterns may reflect self-reinforcing mechanisms, where unfavorable environments depress housing prices, and cheaper areas are favored for pollution-generating infrastructures [14]. Disadvantaged families may then select into these areas due to lower housing costs.

Our findings of higher noise levels increasing the likelihood of excess weight in children contribute to the still limited and conflicting evidence on the relationship between noise exposure and weight in this population. The lack of mediation by sleep duration suggests that qualitative aspects of sleep, such as fragmentation or disturbances, may be more relevant than total duration [8], [9]. Furthermore, differential exposure to environmental stressors did not contribute to the observed socio-economic inequalities in excess weight. These inequalities may instead reflect other factors, such as health behaviors and early-life parenting influences [29]. Conversely, in line with the differential susceptibility hypothesis, we found that the association between air pollution and excess weight tended to be stronger among children from disadvantaged households, although not significantly. This aligns with previous evidence of increased vulnerability to air pollution in disadvantaged adults [14] and echoes findings from a Spanish longitudinal study showing that, at similar residential air pollution levels, deprived children were at greater risk of developing overweight than their advantaged peers [17]. Possible explanations include poorer baseline health and cumulative exposure to other stressors such as second-hand smoke and poor housing quality [17]. Conversely, despite evidence from other studies [10], [11], we did not observe a relationship between nearby greenness and excess weight. Geneva is a relatively safe and green area with 89% of residents living within 300 meters of the nearest green or blue space [26], suggesting that sufficient access to green spaces may reduce barriers to physical and social activities that promote healthy weight.

The absence of associations between urban environmental exposures and excess weight in adolescents was unexpected given previous studies [7], [9], [11]. Compared with children, adolescents might be less affected as they inhale fewer air pollutants per unit of body weight and have greater agency to mitigate noise disturbances [5], [12]. It could also be that adolescents’ school, social, and extracurricular activities take place farther from home than those of children. Residential measures may thus less accurately capture their true environmental exposure, potentially blurring associations.

Overall, the limited associations between urban environmental exposures and excess weight observed in this study may reflect the relatively high quality of Geneva’s urban environment [23], likely driven by effective cantonal policies for a healthy environment [38]. Despite these reassuring findings, our results suggest that childhood weight prevention could benefit from further noise reduction, as well as air quality improvements in disadvantaged areas. This was previously shown in a Chinese study observing that adolescent obesity decreased following environmental policies that improved air quality [39]. Potential measures include redirecting motorized traffic away from residential zones, subsidizing housing insulation, and strengthening pollution controls on vehicles. Moreover, the observed socio-economic disparities in urban environmental exposures warrant attention as they may contribute to inequalities in children’s health beyond excess weight [5]. This is particularly important for policymakers to consider, as current roadmaps for air and green space management do not explicitly take these spatial inequalities into account.

Results of this study should be interpreted in light of several limitations. First, misclassification bias cannot be excluded. Parent-reported weight tends to be underestimated [40]. This bias may be more pronounced in adolescents, as underreporting increases with age [40] and body composition changes rapidly across stages of pubertal maturation, for which we could not adjust. The potentially lower accuracy of excess weight measurement in adolescents might have blurred associations with socio-economic and environmental variables, possibly explaining why such associations were observed in children but not in adolescents. Additionally, several factors that may influence environmental exposure, such as housing insulation, bedroom orientation, and time spent at home were unavailable in our data. Second, the range of environmental exposures was relatively narrow in our study, especially for air pollution (e.g. PM2.5: 7.6-11.4 μg/m^3^), compared with the expected effect size on health (e.g. relative risk of PM2.5 on non-accidental mortality: 1.08 per 10 μg/m^3^)[24], limiting contrast to detect associations. Third, despite random selection of participants, the study sample overrepresented children from highly educated families, when compared to the Geneva population. This imbalance, together with the complete case analysis approach, suggests a possible selection bias. Finally, the cross-sectional design of the study precludes causal inference. Strengths include the population-based design, the wide age range covered, and the ability to examine the independent effects of air and noise pollution and greenness through a careful adjustment strategy.

In conclusion, this study examined the role of multiple urban environmental exposures in socio-economic inequalities in excess weight by testing both the differential exposure and susceptibility hypotheses. We confirmed an association between socio-economic disadvantage and excess weight in children. Differential exposure to air pollution and green areas was observed, while noise was independently related to excess weight in children. However, no environmental exposure explained socio-economic disparities in excess weight. In contrast, we found evidence of differential susceptibility, as the association between air pollution and excess weight tended to be stronger among disadvantaged children. No associations were observed in adolescents, which may reflect lower vulnerability environmental exposures and alternative mechanisms influencing weight outcomes.

Overall, the limited associations observed likely reflect the relatively high quality of the urban environment in Geneva. Nevertheless, specific findings point to the joint influence of social and environmental inequalities on childhood overweight, suggesting that air and noise pollution may represent relevant targets for prevention. Replication in settings with a wider exposure range is warranted to further clarify the mechanisms underlying inequalities in children’s excess weight.

## Data Availability

All data produced in the present study are available upon reasonable request to the authors.

## 5. DECLARATIONS

### 5.1. Ethics approval

The Geneva Cantonal Commission for Research Ethics approved the seroprevalence study (ID: 2020– 00881), the SEROCoV-KIDS study (ID: 2021-01973), and the CIFAR study for further analysis of environmental exposure (ID: 2024-02567).

### 5.3. Authors’ contributions

All authors contributed to the study conceptualization and design. Data curation and methodology were performed by Viviane Richard, David De Ridder, Harris Héritier, Roxane Dumont, Elsa Lorthe, and Nicolas Bovio. Mayssam Nehme, Rémy P. Barbe, Klara M. Posfay-Barbe, Nicolas Vuilleumier, Michael Kobor, Thomas McDade, Idris Guessous and Silvia Stringhini conducted the investigation and funding acquisition. Formal analysis was performed by Viviane Richard who also wrote the original draft of the manuscript. All authors critically reviewed and edited the manuscript. All authors read and approved the final manuscript.

## 5.5. Acknowledgement

We are grateful to the staff of the Unit of Population Epidemiology of the Division of Primary Care Medicine of the University Hospitals of Geneva, as well as to all participants whose contributions were invaluable to the study.

GPT-4o was used in the preparation of this work to improve the readability and language. SEROCoV-KIDS study group: Andrew S. Azman, Antoine Bal, Rémy P. Barbe, Hélène Baysson, Aminata R. Bouhet, Nicolas Bovio, Paola D’Ippolito, Roxane Dumont, Nacira El Merjani, Natalia Fernandez Clares, Natalie Francioli, Idris Guessous, Séverine Harnal, Julien Lamour, Arnaud G L’Huillier, Andrea Loizeau, Elsa Lorthe, Chantal Martinez, Shannon Mechoullam, Mayssam Nehme, Klara M. Posfay-Barbe, Géraldine Poulain, Caroline Pugin, Nick Pullen, Viviane Richard, Deborah Rochat, Khadija Samir, Stephanie Schrempft, Silvia Stringhini, Stéphanie Testini, Deborah Urrutia Rivas, Anshu Uppal, Charlotte Verolet, Jennifer Villers, Guillemette Violot, María-Eugenia Zaballa

