## Supplementary material for "Urban environment and socio-economic inequalities in childhood excess weight: a cross-sectional study in Geneva, Switzerland"

*Supplementary information*

Viviane Richard, David De Ridder, Harris Héritier, Elsa Lorthe, Roxane Dumont, Nicolas Bovio, Mayssam Nehme, Rémy P. Barbe, Klara M. Posfay-Barbe, Thomas W. McDade, Nicolas Vuilleumier, Idris Guessous, Silvia Stringhini, SEROCoV-KIDS study group

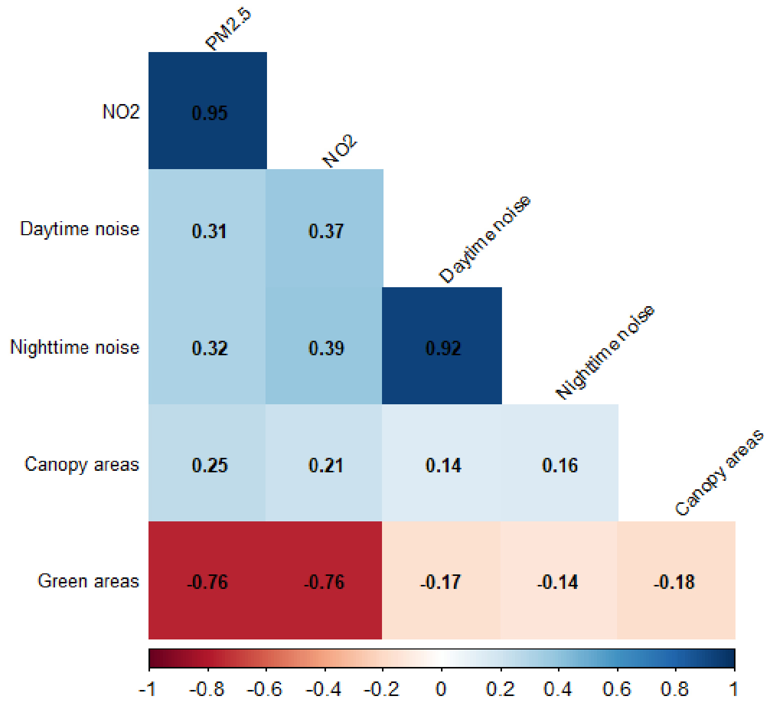


**Supplementary Figure 1.** Pearson correlations coefficients of urban environmental exposures. All corresponding p-values <0.001.

**Supplementary Table 1.** Association between the household financial situation, excess weight and environmental exposures

| | Excess weight | | Air pollution | | Noise pollution | | Greenness | |
|---|---|---|---|---|---|---|---|---|
| | Children (n=923) | Adolescents (n=1'070) | PM2.5 concentration (n=2'008) | $NO_2$ concentration (n=2'008) | Daytime noise (n=2'008) | Nighttime noise (n=2'008) | Canopy area (n=2'008) | Green area (n=2'008) |
| | aOR (95% CI) | aOR (95% CI) | std β (95% CI) | std β (95% CI) | std β (95% CI) | std β (95% CI) | std β (95% CI) | std β (95% CI) |
| **Average to poor household financial situation** (Ref. Good) | 1.79 (1.13; 2.84) | 1.24 (0.82; 1.88) | 0.28 (0.13; 0.43) | 0.25 (0.09;0.40) | 0.07 (-0.06; 0.21) | 0.09 (-0.05; 0.23) | -0.03 (-0.18;0.12) | -0.19 (-0.33; -0.05) |

Results are adjusted odds ratios (aOR), standardized (std) β and 95% confidence intervals (CI) from generalized estimating equations adjusted for age, sex, and parents' birth country. Urban environmental exposures are standardized.

**Supplementary Table 2.** Counterfactual mediation analysis of sleep duration in the association between noise pollution and excess weight in 2-9 years old children (n=921)

| | Sleep duration |
|---|---|
| | aOR (95% CI) |
| **2-9 years old** | |
| *Daytime noise* | |
| Direct effect | 1.35 (1.09-1.69) |
| Indirect/mediating effect | 1.01 (1.00-1.03) |
| Total effect | 1.36 (1.10-1.70) |
| *Nighttime noise* | |
| Direct effect | 1.33 (1.06-1.65) |
| Indirect/mediating effect | 1.01 (1.00-1.03) |
| Total effect | 1.34 (1.07-1.66) |

Results adjusted odds ratios (aOR) and 95% confidence intervals (CI) from counterfactual mediation models, adjusted for age, sex, parents' birth country, household financial situation and types of environmental exposures not included as exposure (i.e. air pollution and greenness measures). Confidence intervals are calculated with bootstraps with 1000 repetitions.

**Supplementary Table 3.** Moderating effect of the household financial situation in the association of urban environmental exposures with excess weight in 2-9 (n=923) and 10-17 (n=1'070) years old

| | Excess weight |
|---|---|
| | aOR (95% CI) |
| *2-9 years* | |
| **Air pollution** | |
| PM2.5 concentration | 1.59 (0.98; 2.59) |
| NO2 concentration | 1.47 (0.92; 2.35) |
| **Noise pollution** | |
| Daytime noise | 1.08 (0.66; 1.74) |
| Nighttime noise | 1.12 (0.70; 1.79) |
| **Greenness** | |
| Canopy area | 1.01 (0.66; 1.56) |
| Green area | 0.76 (0.46; 1.26) |
| *10-17 years* | |
| **Air pollution** | |
| PM2.5 concentration | 0.84 (0.55; 1.27) |
| NO2 concentration | 0.84 (0.55; 1.27) |
| **Noise pollution** | |
| Daytime noise | 1.06 (0.71; 1.59) |
| Nighttime noise | 1.11 (0.75; 1.64) |
| **Greenness** | |
| Canopy area | 1.13 (0.71; 1.79) |
| Green area | 1.40 (0.90; 2.16) |

Results are the coefficient of the interaction between the household financial situation and urban environmental exposures expressed as adjusted odds ratios (aOR) and 95% confidence intervals (CI) from generalized estimating equations. Models are adjusted for age, sex, parents' birth country and types of environmental exposures not included as exposure (e.g. the association between canopy area and excess weight is adjusted for air and noise pollution measures). Urban environmental exposures are standardized; coefficients express the odds of excess weight for a change of one standard deviation in the environmental exposure in those with an average to poor (versus good) household financial situation.
